# Comparison of Evaluation Metrics of Deep Learning for Imbalanced Imaging Data in Osteoarthritis Studies

**DOI:** 10.1101/2022.09.08.22279696

**Authors:** Shen Liu, Frank Roemer, Yong Ge, Edward J. Bedrick, Zong-Ming Li, Ali Guermazi, C. Kent Kwoh, Xiaoxiao Sun

## Abstract

**Objective:** To compare the evaluation metrics for deep learning methods in the imbalanced imaging data in osteoarthritis (OA) studies.

**Method:** We first divided MOAKS (MRI Osteoarthritis Knee Score) grades into the presence (MOAKS > 0) and absence (MOAKS = 0) categories. Second, a deep-learning model was trained to the sagittal intermediate-weighted (IW) fat-suppressed (FS) knee MRI images with MOAKS readings from the Osteoarthritis Initiative (OAI) study to predict the presence of bone marrow lesions (BMLs). After the deep learning models were trained, we obtained probabilities of the presence of BMLs from MRI images at the sub-region (15 sub-regions), compartment, and whole-knee levels. We compared different evaluation metrics (e.g., receiver operating characteristic (ROC) and precision-recall (PR) curves) of the deep learning model in the testing data with various class ratios (presence of BMLs vs. absence of BMLs) at these three data levels to assess the model’s performance.

**Results:** We have demonstrated that the commonly used ROC curve is not sufficiently informative when evaluating the performance of deep learning models in the imbalanced data in OA studies.

**Conclusion:** The class ratios coupled with results of ROC, PR, and Matthews correlation coefficient (MCC) should be reported in OA studies.

## 1. Introduction

Knee osteoarthritis (OA) is a leading cause of impaired mobility and disability in the aging population^1^, affecting more than 50 million people in the United States ^2^. As there are no effective therapies to slow or stop the progression of knee OA^3^, the only effective treatment for late-stage knee OA is total knee replacement (TKR)^4^. The number of TKRs is projected to be more than 3 million in 2030^5^. Therefore, preventive efforts and interventions earlier in the disease course could significantly reduce the number of TKR procedures and thus greatly reduce the public health impact of knee OA.

Prior studies have shown that some OA features only detectable by MRI, including bone marrow lesions (BMLs) play a role in structural OA incidence and progression and may be present before cartilage damage occurs^6–8^. BMLs are associated with knee pain and meniscal damage and extrusion^9,10^. One approach for detailed characterization of BMLs is based on ordinal expert grading using semi-quantitative (SQ) scoring systems (e.g., MRI Osteoarthritis Knee Score (MOAKS)) on MRI^13^. Application of SQ scoring systems for structural tissue characterization requires experienced and trained musculoskeletal radiologists to perform these MRI readings, which is both time-consuming and resource-intensive.

Deep learning (DL) methods (e.g., convolutional neural network (CNN) models^14^) have been implemented to automatically detect MRI-defined abnormalities from MRI ^15–17^. The area under the receiver operating characteristic (ROC) curve^18^ is usually utilized to assess the performance of a DL-based binary classifier for classifying positive (presence of MRI-defined features) and negative classes. The ROC curve plots the true positive rates against the false positive rates given various threshold settings. The area under the ROC curve (ROC-AUC) ranges from 0 to 1, with 1 indicating perfect diagnostic performance. In general, a binary classifier with a ROC-AUC value of 0.8 to 0.9 is considered excellent and has an outstanding performance with a value of more than 0.9. However, ROC-AUC alone is not sufficiently informative to evaluate the performance of approaches when the underlying data have class imbalance problems (i.e., positive class occurs with a markedly reduced frequency) as ROC-AUC is insensitive to the changes in class imbalance ratios in the data, resulting in uninformative evaluation of model performance^19,20^. Since most of the datasets from large-scale OA studies are imbalanced with many features being rare ^16,21–25^, it is important to consider frequencies of findings in order to define whether a given dataset is balanced and/or imbalanced when utilizing DL methods.

Aim of this study was to compare several metrics for evaluating the performance of DL-based models using data with various class imbalance ratios. To perform this comparison, we implemented and modified an existing deep learning framework^25^ to predict the presence of BMLs using MRIs from the Osteoarthritis Initiative (OAI) study.

## 2. Materials and Method

### 2.1. Datasets

We used all MRIs of the OAI with available MOAKS readings at baseline. The OAI is a large longitudinal cohort study of participants with or at risk of developing knee OA including MRI data^26^. 4,796 participants were enrolled at four sites: Ohio State University, University of Maryland/Johns Hopkins University, Memorial Hospital (Providence, RI), and the University of Pittsburgh. Bilateral knee MRI scans were obtained at baseline and over the following eight years using identical 3T MR systems (Siemens MAGNETOM Trio, Erlangen, Germany). The clinical and imaging data in the OAI are publicly available. Among 4,796 participants, MRIs of 2,473 subjects were assessed by radiologists using the semi-quantitative scoring system (i.e., MOAKS). After excluding 6 participants without MOAKS readings at baseline, the sagittal intermediate-weighted (IW fat-suppressed (FS) MRI sequence of 2,467 participants were utilized for the data analysis. Subchondral bone marrow signal alterations are characterized by ill-defined subchondral areas of high signal intensity IW FS MRIs^8,27^. The knee joint is subdivided into 15 subregions for BML assessment (Supplementary Table 1) in the MOAKS scoring system. We used the BML size score, which is the percentage of volume per subregion including associated cystic BMLs^13^. The dataset was split into a training set (1838 exams, 1500 subjects), a validation set (582 exams, 480 subjects), and a testing set (576 exams, 487 subjects) by random sampling. Among all the sagittal IW FS sequences, the number of slices included varied from 34 to 43 (mean is 37.30 with a standard deviation of 0.85).

### 2.2 Preprocessing

The input images were extracted from Digital Imaging and Communications in Medicine (DICOM) files, scaled to 256 × 256 pixels, normalized into the intensity values between 0 and 255, and converted to the Portable Network Graphics (PNG) format using the pydicom (version 0.9.9), nibabel (version 3.2.1) and nyul (version 1.3) libraries from Python (version 3.6.5). The histogram-based intensity standardization algorithm^28,29^ was utilized to standardize the PNG images to improve image similarities between scans and compare images from different scans fairly. In particular, the distribution of intensity values of all images in the training set was learned to linearly map the intensity values of the images of testing and validation sets.

## Methods

### Deep learning framework

We applied a deep learning framework (MRNet^25^) to the preprocessed MRIs. The convolutional neural network (CNN) is used in MRNet to map the MRIs to the probabilities of the presence/absence of BMLs. The input of MRNet is the slices of each scan with the dimension of ! × 3 × 256 × 256, where ! is the number of slices in this scan. If the MRI scan was from the right knee, the first half of MRI slices were selected into the model for detecting BMLs in sub-regions of the lateral side; and the second half of MRI slices were used for predicting BMLs in sub-regions of the medial side. If the MRI scan was from the left knee, the half of slices selected into the model were opposite to those in the MRIs from the right knee described above. The binary cross-entropy loss function was minimized to optimize the deep learning framework. We dichotomized the MOAKS grades into presence or absence categories. The split was done by categorizing grades > 0 as presence (i.e., positive class) and grades = 0 as absence (i.e., negative class). The prevalence of BMLs in each of the training, validation, and testing dataset at the whole-knee, compartment, and sub-region levels is shown in Supplementary Table 2. The images with the dichotomized MOAKS grades were used in the model training process. We implemented backpropagation algorithm to optimize the deep learning model. To avoid over-fitting issues, we rotated (−10 to 10 degrees) and shifted (−25 to 25 pixels) the images randomly. The model with the smallest average loss values on the validation set was selected as the final model for prediction. With an NVIDIA V100S 32GB GPU, the training process took 1.5 hours for ten iterations. The MRNet framework was executed with Python (version 3.6.5) and PyTorch (version 1.2.0).

### Model diagnostics

We used generated class activation mappings (CAMs^31^) to highlight the regions contributing to the prediction of BMLs. The highlighted regions allow researchers to diagnose the fitted deep learning models by inspecting if the highlighted regions contain BMLs. In particular, the weights from the classification layers in neural networks were obtained to compute a weighted average CNN feature map, which was mapped to the original image with a color scheme after up-sampling.

### Whole-knee level, compartment-level and sub-region level prediction

We obtained predicted BML status (presence vs. absence) for each of the 15 sub-regions from the trained DL models. With the above prediction of 15 sub-regions as independent variables and overall MOAKS score as the dependent variable, which was dichotomized as 0 (all 15 sub-regions have MOAKS readings of 0) and 1 (at least one of 15 sub-regions has MOAKS reading of 1), a logistic regression model was implemented to predict BML status at the whole-knee level. The compartment-level prediction is similar to the whole-knee level prediction procedure. This prediction was performed by the scikit-learning (version 0.24.1) package in Python (version 3.6.5).

### Performance evaluation

The evaluation metrics of DL-based methods for binary classification are based on the confusion matrix, a contingency table to summarize the performance of models (i.e., true positive (‘(), false negative ()*), false positive ()(), and true negative (‘*)). The ROC curve plots the true positive rate (‘(/(‘(+)*)) against the false positive rate ()(/()(+ ‘*)) for different threshold values, which can be used to assign observations to the positive class when the scores (e.g., probabilities of the presence of BMLs) from the model are above a threshed value. The precision-recall (PR) curve^32^ depicts the relationship between recall (x-axis) and precision (y-axis). The precision-recall gain (PRG) curve^33^ plots the precision gain against recall gain, where the precision and recall gains are adjusted by the proportion of positives. The maximum value of the AUC of the ROC, PR, and PRG curves is one, showing perfect prediction performance. Notice that the PRG curve is adjusted by the class ratio. We also consider two metrics that can be expressed as single numbers for the fixed threshold value (e.g., default threshold values). The F1^34^ score is defined as the harmonic mean of precision and recall. The Matthews correlation coefficient^35^ (MCC) measures the Pearson product-moment correlation coefficient between the true and predicted instances and is usually not affected by the class imbalance issue. The worst and best values of MCC are -1 and +1, respectively. The specificity is defined as the true negative rate. We compared these metrics in the testing set.

### Strategies to address the imbalanced data

Several DL-based methods have been developed for imbalanced data. For instance, a new loss function^37–39^ that is sensitive to the errors from the positive class and a data-level method^40^ that can down-sample the negative class can be applied. We applied this down-sample method to our data. We also removed the weights in the loss function of MRNet to examine the importance of the weights for the imbalanced data.

## 3. Results

The baseline demographic statistics of selected participants are shown in Table 1. A majority of subjects (i.e., 56.67%-61.67% in different datasets) were female. The mean age and BMI were approximately 61 years and 28, respectively. The mean differences in terms of age and BMI across all datasets (i.e., training, validation, and testing) were not statistically significant. In Table 2, we show the results of different performance metrics at the whole-knee, compartment, and sub-region levels. At the whole-knee level, the ROC-AUC value was 0.86, indicating excellent performance. The model also had a PR-AUC value of 0.96, an F1 score of 0.88, a sensitivity value of 0.88, and a precision value of 0.86. However, a different conclusion about the model performance at the whole-knee level could be drawn according to the values of PRG-AUC (0.42) and MCC (0.36). As shown in Figure 1A, the class ratio (positive/negative) for the whole-knee level data was 0.79/0.21. If BMLs were present in one of the 15 sub-regions, the class label for this knee was positive. Thus, the negative class was the minority class in the whole-knee level imbalanced data. The specificity value was 0.43, indicating the ability to predict negatives was limited for the whole-knee model, which was also confirmed by the MCC value. Since the PRG curve is adjusted by the proportion of positives, its AUC value might be affected when the data are imbalanced (i.e., the proportion of positives is close to zero or one). As shown in Figure 2A, the PRG curve had the smallest AUC value.

**Table 1.**
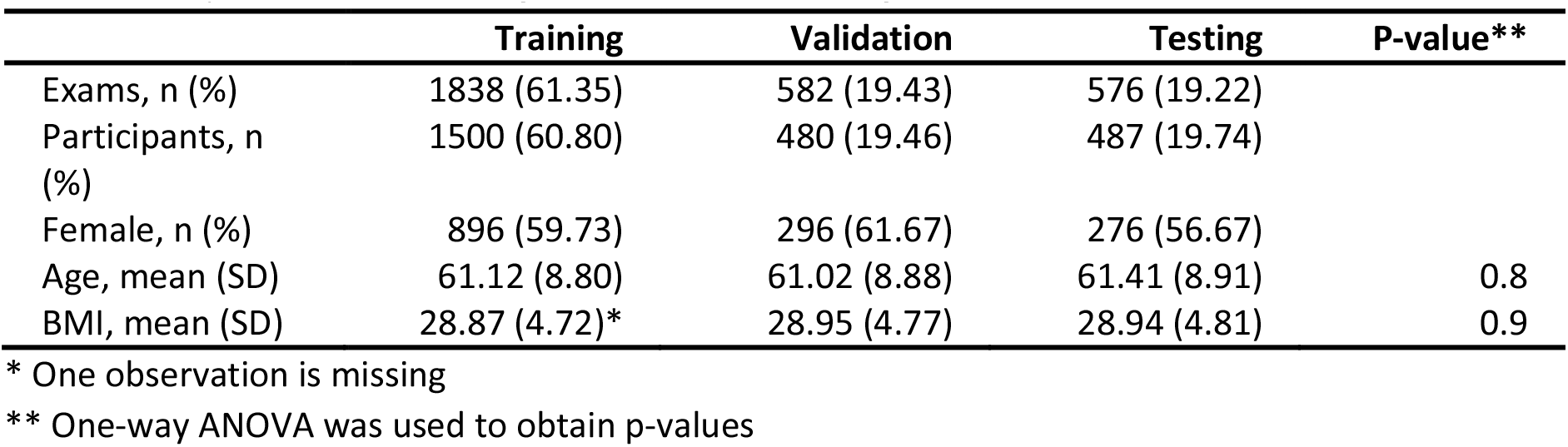
Summary statistics of training, validation, and testing datasets

**Table 2.** Performance evaluation at the whole-knee, compartment, and sub-region levels

|  | ROC-AUC | PR-AUC | PRG-AUC | F1 | MCC | Precision | Sensitivity (Recall) | Specificity |
| --- | --- | --- | --- | --- | --- | --- | --- | --- |
| Whole-knee | 0.86 | 0.96 | 0.42 | 0.88 | 0.36 | 0.86 | 0.90 | 0.43 |
| Medial | 0.73 | 0.76 | 0.46 | 0.66 | 0.34 | 0.65 | 0.66 | 0.68 |
| Lateral | 0.87 | 0.80 | 0.89 | 0.71 | 0.61 | 0.78 | 0.65 | 0.92 |
| Patella | 0.87 | 0.89 | 0.80 | 0.77 | 0.54 | 0.74 | 0.81 | 0.73 |
| FemAntLat | 0.95 | 0.85 | 0.98 | 0.61 | 0.55 | 0.45 | 0.95 | 0.74 |
| FemAntMed | 0.66 | 0.32 | 0.46 | 0.12 | 0.11 | 0.45 | 0.07 | 0.98 |
| FemCentLat | 0.92 | 0.63 | 0.99 | 0.26 | 0.38 | 1 | 0.15 | 1 |
| FemCentMed | 0.91 | 0.82 | 0.95 | 0.74 | 0.68 | 0.82 | 0.67 | 0.96 |
| FemPostLat | 0.96 | 0.79 | 1 | 0.67 | 0.68 | 0.91 | 0.53 | 1 |
| FemPostMed | 0.79 | 0.36 | 0.8 | 0.34 | 0.28 | 0.44 | 0.27 | 0.95 |
| TibSubSpCent | 0.87 | 0.76 | 0.87 | 0.67 | 0.55 | 0.72 | 0.62 | 0.90 |
| TibAntLat | 0.84 | 0.10 | 0.76 | 0 | 0 | 0 | 0 | 1 |
| TibAntMed | 0.89 | 0.63 | 0.96 | 0.62 | 0.57 | 0.65 | 0.59 | 0.96 |
| TibCentLat | 0.86 | 0.56 | 0.96 | 0.54 | 0.49 | 0.5 | 0.59 | 0.94 |
| TibCentMed | 0.94 | 0.85 | 0.97 | 0.78 | 0.73 | 0.74 | 0.83 | 0.94 |
| TibPostLat | 0.76 | 0.28 | 0.9 | 0.37 | 0.33 | 0.42 | 0.33 | 0.97 |
| TibPostMed | 0.84 | 0.33 | 0.88 | 0.14 | 0.15 | 0.38 | 0.08 | 0.99 |
| PatellaLat | 0.88 | 0.78 | 0.9 | 0.70 | 0.59 | 0.66 | 0.75 | 0.86 |
| PatellaMed | 0.54 | 0.38 | 0.1 | 0.03 | 0.07 | 0.75 | 0.02 | 1 |

**Figure 1.**
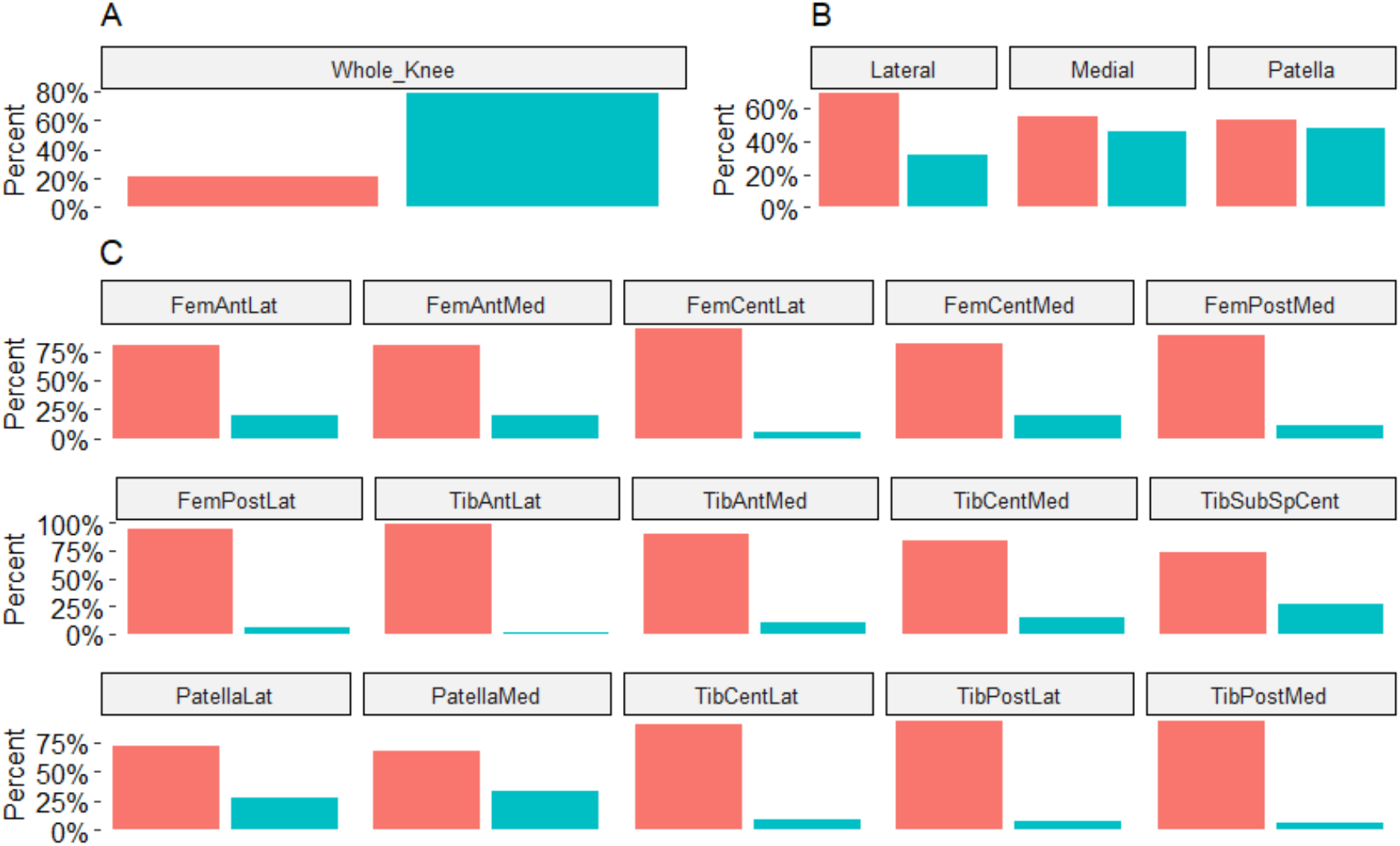
Class ratios (negative (red) : positive (cyan)) at A) subject level; B) compartment level; C) sub-region level

**Figure 2.**
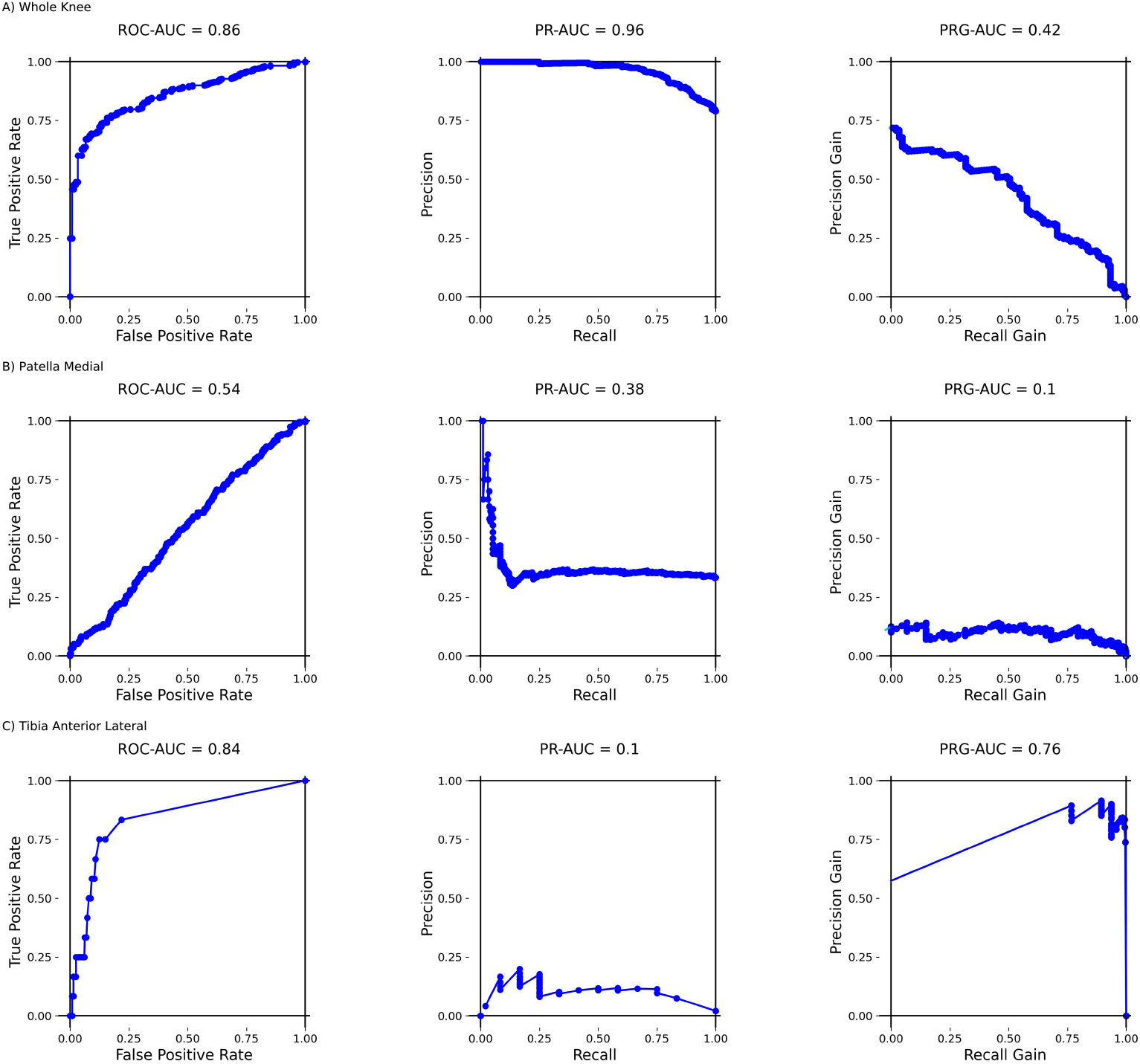
ROC (left panels), PR (middle panels), and PRG (right panels) curves at A) whole-knee level; B) *PatellaMed*; C) *TibAntLat*. The AUC values of the curves are shown on top of each figure.

At the compartment level, there was a mild class imbalance issue (i.e., about 60% negative) in the lateral compartment (see Figure 1B). Although the class ratio was around 1:2 for the lateral femorotibial (TF) compartment, there were still a sufficient number of positives to train and tune the DL model. The class ratios were approximately 1:1 for the medial TF and patellofemoral (PF) compartments. Therefore, all metrics presented consistent information about the model performance for each compartment. For instance, the DL model obtained better performance in the lateral TF and PF compartments than in the medial TF compartment.

At the sub-region level, data were severely imbalanced, with less than 10% positives in some sub-regions (see Figure 1C). The abbreviation and list of these subregions are shown in Supplementary Table 1. The PR-AUC and MCC had the highest correlation value (0.92), whereas the correlation value between ROC-AUC and MCC was 0.74. The metrics F1 and MCC had consistent conclusions in terms of model performance, with a correlation value of 0.98. The ROC-AUC values of the *FemAntMed, FemPostMed, TibPostLat*, and *PatellaMed* sub-regions were below 0.8. The results of these metrics (i.e., MCC, F1, PR-AUC) were consistent with those of ROC-AUC. However, PRG-AUC for the *TibPostLat* sub-region (i.e., 0.9) was too high due to its dependence on the proportion of positives. Based on the value of MCC, the prediction performance for this sub-region might not be considered as outstanding. The ROC-AUC values of the rest sub-regions were above 0.8. For example, for the *PatellaMed* sub-region, the classification results were not as good as those in other sub-regions. The ROC, PR, and PRG curves provided complementary information, see Figure 2B. The ROC-AUC value was 0.84 in the *TibAntLat* sub-region, showing excellent prediction performance; nevertheless, the value of PR-AUC was only 0.10 in the *TibAntLat* sub-region (see Figure 2C). For this sub-region, PR-AUC was more informative since the precision and sensitivity were zeros, indicating that all data were assigned to the negative class. The MCC and F1 metrics were also good indicators for the performance evaluation for classifying the positive class, with the values of zeros. This might be due to the high imbalance ratio (46 positives:1223 negatives) in the data for the *TibAntLat* sub-region (see Supplementary Table 2).

The down-sampling method improved the performance of DL methods under an imbalanced scenario (see Supplementary Table 3). Such an improvement was trivial in the sub-regions such as *TibAntLat*, indicating that the original class ratios were critical for model training. The weights in the loss function are important for the imbalanced data^36^. Without these weights in MRNet, MCC became zeros in some sub-regions (e.g., *FemAntMed* and *TibCentMed*) (see Supplementary Table 4).

Recently, CAM^31^ is becoming the standard method to interpret the prediction decisions and perform model diagnostics of DL models. Suppose there are not enough positives or negatives to train and tune the DL models. In those instances, the highlighted regions generated by CAM may not be overlapping with the regions of MRI-defined abnormalities (e.g., BMLs) (see Figure 3). Such an approach is useful to evaluate the results in DL models and should also be reported.

**Figure 3.**
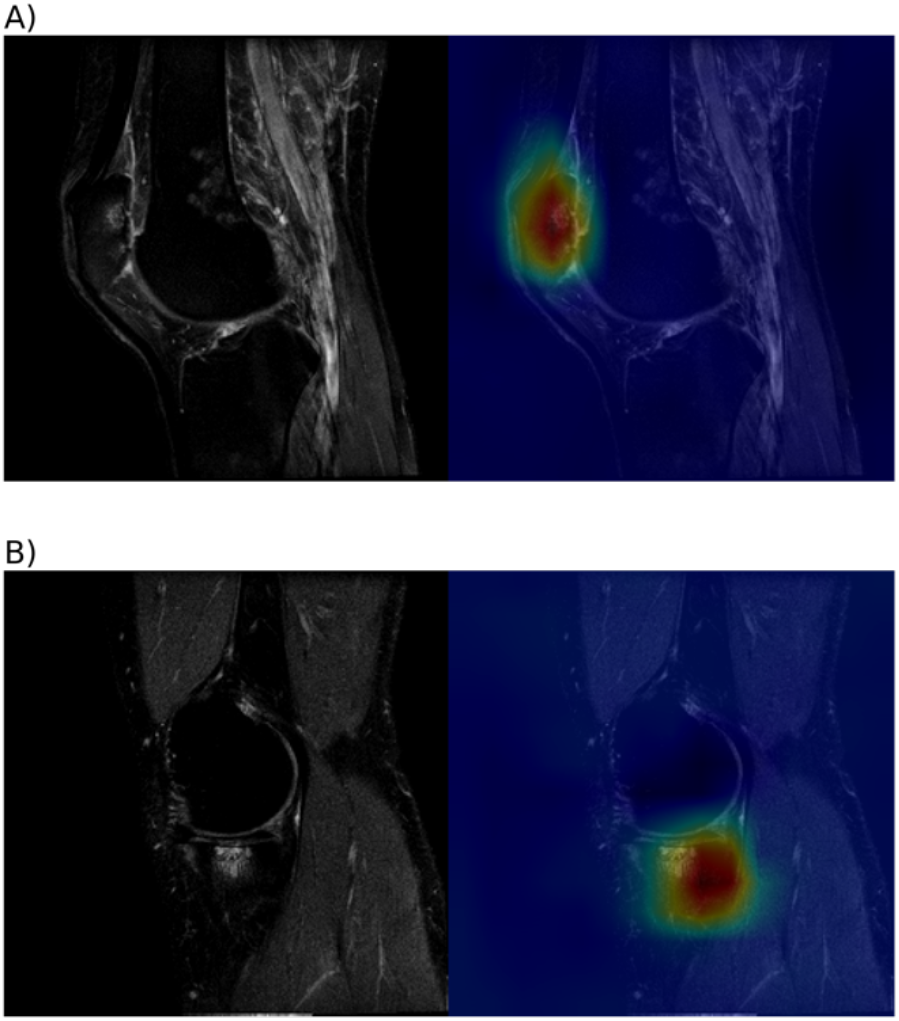
A) The original MRI image is on the left-hand side (BMLs on *Lateral* side of Patella); CAM mappings (highlighted regions) are on the right-hand side. B) The original MRI image is on the left-hand side (BMLs on *Medial* side of Tibia); CAM mappings (highlighted regions) are on the right-hand side.

We also checked the distribution of all MOAKS grades (i.e., 0-3) at the sub-region level and found it was highly skewed/imbalanced (see Supplementary Figure 1). In some sub-regions (e.g., *TibAntLat*), there were no MOAKS grades of 2 and 3, which was challenging to train the non-binary DL models to predict these grades. Furthermore, in most deep learning applications, all data are typically spitted into three datasets (i.e., training, validation, and testing) by random sampling. This subsampling process may also aggravate the class imbalance concerns (see Supplementary Figures 2, 3, and 4). For example, there was no MOAKS grade 3 in *TibCentLat* in the validation data to tune and/or optimize the DL models. We also examined the class imbalance problems in the sub-studies (e.g., FNIH). The data were also severely imbalanced, as shown in Supplementary Table 5. For instance, in the *FemCentLat* and *FemPostLat* sub-regions, there were less than ten positives in training, validation, and testing datasets.

## 4. Discussion

Osteoarthritis is a multifactorial disease of the whole joint and may result in substantial pain and poor quality of life in patients. Recently, numerous DL methods have been applied to automatically detect different MRI-defined abnormalities in OA. The development and validation of accurate DL frameworks to detect MRI-defined abnormalities are critical for the early detection of OA and optimal participant screening for clinical trials of disease-modifying OA drugs (DMOAD). The ultimate application of these DL models is dependent on informative reporting of the performance metrics used to benchmark different DL models, particularly when the underlying MRIs used to derive these models are imbalanced^16,21–25^. To compare the performance metrics of DL methods for imbalanced and/or balanced MRI data, we have modified an existing deep learning framework (MRNet) to detect BMLs from the MRI scans automatically.

Our findings suggest the class imbalance ratios can affect the evaluation results of some metrics since the DL learners may over-classify the majority group (i.e., assign all minorities to majorities) to obtain high overall accuracy. In the OAI study, the severity of class imbalance for BMLs was associated with the data levels (i.e., whole-knee, compartment, and sub-region levels). Specifically, at the whole-knee and sub-region levels, the class imbalance issues were severe, while the data were balanced at the compartment level. When data were balanced at the compartment level, all the evaluation metrics had consistent results. With the imbalanced data at the whole-knee and sub-region levels, the commonly used metric ROC-AUC alone might provide a distorted view of the DL methods, demonstrating excellent performance (> 0.8) even for the classifiers that assign all positives to negatives. The ROC curve is necessary but not sufficient to evaluate the performance of DL methods, particularly when data are imbalanced^36^. In our case study, the PR curve can be used to address the issues due to the class imbalance. The PRG curve is overly optimistic when data are severely imbalanced. Therefore, multiple metrics, including ROC-AUC, PR-AUC, F1, and MCC, are highly recommended for evaluating model performance. The ROC and PR curves are rank metrics under different threshold settings, while F1 and MCC are threshold metrics for a fixed threshold setting. The ordering of prediction instead of the actual predicted values is used in rank metrics, while the threshold metric depends on a threshold level (e.g., data are predicted as positives above the threshold level). Since the rank metrics provide a summary of model performance for all possible threshold settings, the ROC-AUC and PR-AUC can be used as the major evaluation metrics. The F1 and MCC can be used as the auxiliary metrics to evaluate model performance. In addition, the class imbalance ratio should be reported.

We applied several methods to address the underlying imbalanced issues. Although the performance of DL methods improved, in general, the amount of improvement was not clinically significant. These results indicate that the underlying imbalance reflected by the class ratios was too severe to overcome by these methods. The DL algorithms have been widely used in detecting various features of OA (e.g., effusion-synovitis and osteophytes) using the medical imaging data. Our findings can be generalized to the studies using any radiologic imaging data with the class imbalanced issues. In the present work, we considered only the binary cases (i.e., BMLs vs. non-BMLs). The comparison of performance metrics for the multi-class classifiers (e.g., MOAKS grades 0-3) is an area for future research.

## Data Availability

All data produced in the present study are available upon reasonable request to the authors

## 5. Contributions

Conception and design: XS, KK, SL Image Analysis: SL, XS Statistical Analysis: SL, XS Drafting of Article: SL, XS, KK, YG Review/revision: XS, KK, FWR, YG, AG, EB, ZL

---

**Supplementary Table 1.**
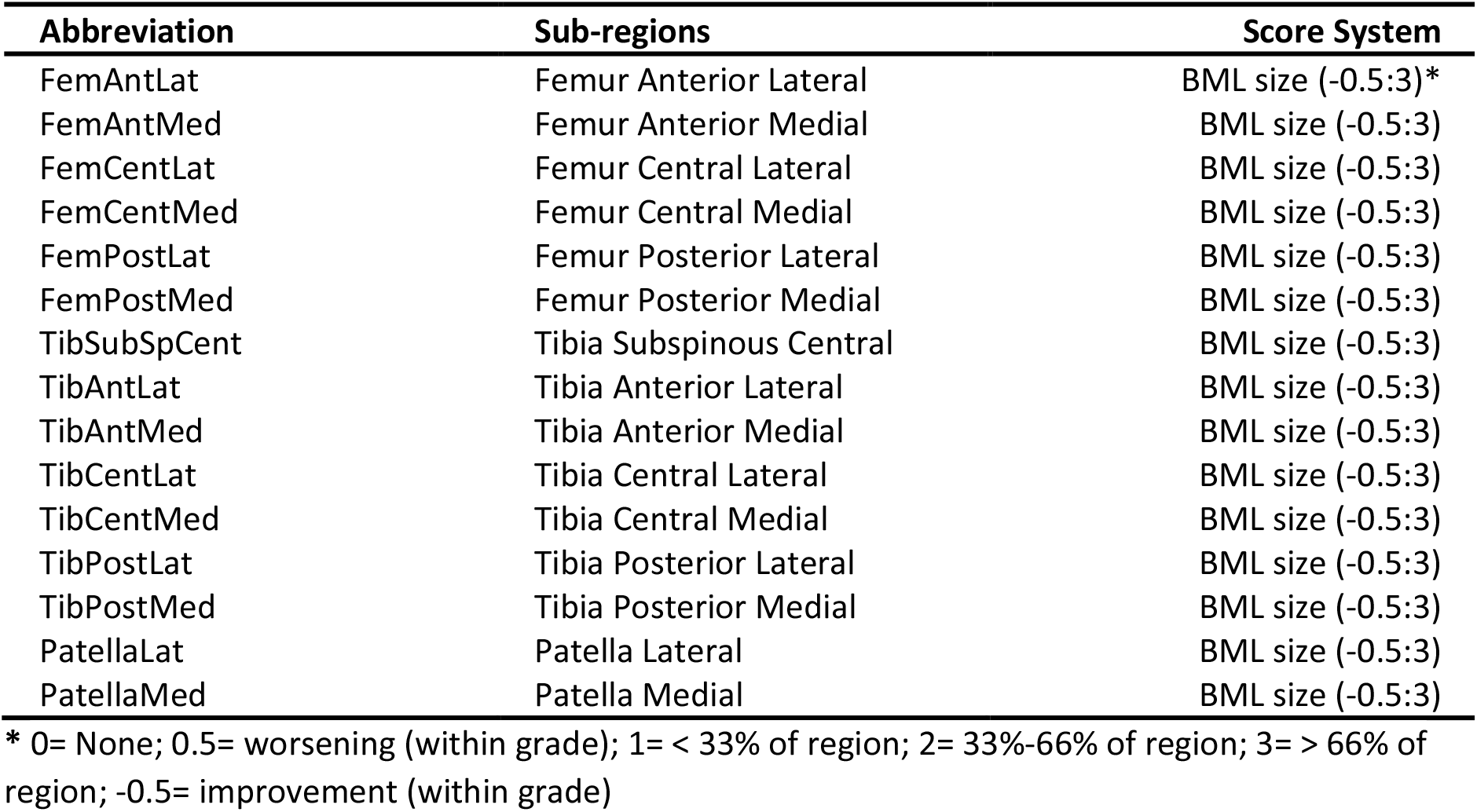
Sub-regions and score systems considered in this study

**Supplementary Table 2.** Class ratios (positive:negative) of training, validation and testing datasets at the whole-knee, compartment and sub-region levels

|  | Total | Training | Validation | Testing |
| --- | --- | --- | --- | --- |
| Whole-knee | 2345:651 | 1440:398 | 450:132 | 455:121 |
| Medial | 1360:1636 | 836:1002 | 250:332 | 274:302 |
| Lateral | 945:2051 | 594:1244 | 183:399 | 148:408 |
| Patella | 1419:1577 | 849:989 | 288:294 | 282:294 |
| FemAntLat | 580:2416 | 363:1475 | 112:470 | 105:471 |
| FemAntMed | 609:2387 | 361:1477 | 123:459 | 125:451 |
| FemCentLat | 162:2834 | 107:1731 | 28:554 | 27:549 |
| FemCentMed | 569:2427 | 348:1490 | 95:487 | 126:450 |
| FemPostLat | 169:2827 | 114:1724 | 36:546 | 19:557 |
| FemPostMed | 324:2672 | 200:1638 | 54:528 | 70:506 |
| TibSubSpCent | 819:2177 | 487:1351 | 162:420 | 170:406 |
| TibAntLat | 49:2947 | 32:1806 | 5:577 | 12:564 |
| TibAntMed | 303:2693 | 183:1655 | 54:528 | 66:510 |
| TibCentLat | 269:2727 | 171:1667 | 44:538 | 54:522 |
| TibCentMed | 462:2534 | 273:1565 | 90:492 | 99:477 |
| TibPostLat | 211:2785 | 123:1715 | 49:533 | 39:537 |
| TibPostMed | 191:2805 | 115:1723 | 40:542 | 36:540 |
| PatellaLat | 845:2151 | 503:1335 | 189:393 | 153:423 |
| PatellaMed | 982:2014 | 588:1250 | 202:380 | 192:384 |

**Supplementary Table 3.** Performance evaluation after down sampling at the sub-region level

|  | ROC-AUC | PR-AUC | PRG-AUC | Precision | Sensitivity (Recall) | Specificity | F1 | MCC |
| --- | --- | --- | --- | --- | --- | --- | --- | --- |
| FemAntLat | 0.93 | 0.78 | 0.96 | 0.37 | 0.96 | 0.63 | 0.53 | 0.46 |
| FemAntMed | 0.59 | 0.30 | 0.29 | 0.27 | 0.55 | 0.60 | 0.37 | 0.12 |
| FemCentLat | 0.91 | 0.47 | 0.98 | 0.31 | 0.70 | 0.92 | 0.43 | 0.43 |
| FemCentMed | 0.88 | 0.74 | 0.91 | 0.52 | 0.77 | 0.80 | 0.62 | 0.50 |
| FemPostLat | 0.89 | 0.25 | 0.88 | 0.11 | 0.89 | 0.76 | 0.20 | 0.27 |
| FemPostMed | 0.73 | 0.27 | 0.65 | 0.24 | 0.53 | 0.77 | 0.33 | 0.22 |
| TibSubSpCent | 0.83 | 0.72 | 0.82 | 0.50 | 0.84 | 0.65 | 0.63 | 0.45 |
| TibAntLat | 0.88 | 0.10 | 0.76 | 0.09 | 0.42 | 0.91 | 0.15 | 0.16 |
| TibAntMed | 0.83 | 0.48 | 0.91 | 0.26 | 0.73 | 0.73 | 0.38 | 0.31 |
| TibCentLat | 0.76 | 0.32 | 0.85 | 0.26 | 0.54 | 0.84 | 0.35 | 0.28 |
| TibCentMed | 0.91 | 0.75 | 0.96 | 0.47 | 0.85 | 0.80 | 0.60 | 0.53 |
| TibPostLat | 0.69 | 0.20 | 0.79 | 0.38 | 0.15 | 0.98 | 0.22 | 0.21 |
| TibPostMed | 0.83 | 0.25 | 0.89 | 0.14 | 0.83 | 0.67 | 0.25 | 0.26 |
| PatellaLat | 0.69 | 0.44 | 0.52 | 0.35 | 0.81 | 0.46 | 0.49 | 0.24 |
| PatellaMed | 0.54 | 0.38 | 0.09 | 0.34 | 0.91 | 0.13 | 0.50 | 0.06 |

**Supplementary Table 4.** Performance evaluation of MRNet without weights in the loss function at the sub-region level

|  | ROC-AUC | PR-AUC | PRG-AUC | Precision | Sensitivity (Recall) | Specificity | F1 | MCC |
| --- | --- | --- | --- | --- | --- | --- | --- | --- |
| FemAntLat | 0.96 | 0.87 | 0.98 | 0.90 | 0.70 | 0.98 | 0.79 | 0.76 |
| FemAntMed | 0.57 | 0.26 | 0.23 | 0 | 0 | 1 | 0 | 0 |
| FemCentLat | 0.93 | 0.60 | 0.97 | 0.89 | 0.30 | 1 | 0.44 | 0.50 |
| FemCentMed | 0.83 | 0.71 | 0.90 | 0.82 | 0.97 | 0.82 | 0.60 | 0.55 |
| FemPostLat | 0.93 | 0.72 | 0.99 | 0.85 | 0.58 | 1 | 0.69 | 0.69 |
| FemPostMed | 0.76 | 0.33 | 0.74 | 0.50 | 0.09 | 0.99 | 0.15 | 0.17 |
| TibSubSpCent | 0.85 | 0.75 | 0.85 | 0.63 | 0.72 | 0.82 | 0.67 | 0.52 |
| TibAntLat | 0.85 | 0.07 | 0.47 | 0 | 0 | 1 | 0 | 0 |
| TibAntMed | 0.89 | 0.60 | 0.96 | 0.82 | 0.27 | 0.99 | 0.41 | 0.44 |
| TibCentLat | 0.76 | 0.34 | 0.87 | 0 | 0 | 1 | 0 | 0 |
| TibCentMed | 0.95 | 0.83 | 0.97 | 0.82 | 0.66 | 0.97 | 0.73 | 0.69 |
| TibPostLat | 0.68 | 0.15 | 0.73 | 0 | 0 | 1 | 0 | 0 |
| TibPostMed | 0.83 | 0.33 | 0.91 | 0.38 | 0.44 | 0.95 | 0.41 | 0.36 |
| PatellaLat | 0.82 | 0.65 | 0.79 | 0.54 | 0.71 | 0.78 | 0.61 | 0.45 |
| PatellaMed | 0.54 | 0.36 | 0.09 | 0 | 0 | 1 | 0 | 0 |

**Supplementary Table 5.** Class ratios (positive:negative) of training, validation and testing datasets in the FNIH study at the sub-region level

|  | Total | Training | Validation | Testing |
| --- | --- | --- | --- | --- |
| FemAntLat | 136:314 | 87:180 | 22:59 | 27:75 |
| FemAntMed | 124:326 | 66:201 | 25:56 | 33:69 |
| FemCentLat | 16:434 | 9:258 | 6:75 | 1:101 |
| FemCentMed | 126:324 | 70:197 | 23:58 | 33:69 |
| FemPostLat | 12:438 | 10:257 | 1:80 | 1:101 |
| FemPostMed | 76:374 | 41:226 | 12:69 | 23:79 |
| TibSubSpCent | 138:312 | 84:183 | 19:62 | 35:67 |
| TibAntLat | 2:448 | 2:265 | 0:81 | 0:102 |
| TibAntMed | 63:387 | 35:232 | 10:71 | 18:84 |
| TibCentLat | 24:426 | 17:250 | 3:78 | 4:98 |
| TibCentMed | 110:340 | 62:205 | 20:61 | 28:74 |
| TibPostLat | 26:424 | 14:253 | 7:74 | 5:97 |
| TibPostMed | 30:420 | 19:248 | 7:74 | 4:98 |
| PatellaLat | 162:288 | 103:164 | 31:50 | 28:74 |
| PatellaMed | 154:296 | 90:177 | 24:57 | 40:62 |

**Supplementary Figure 1.**
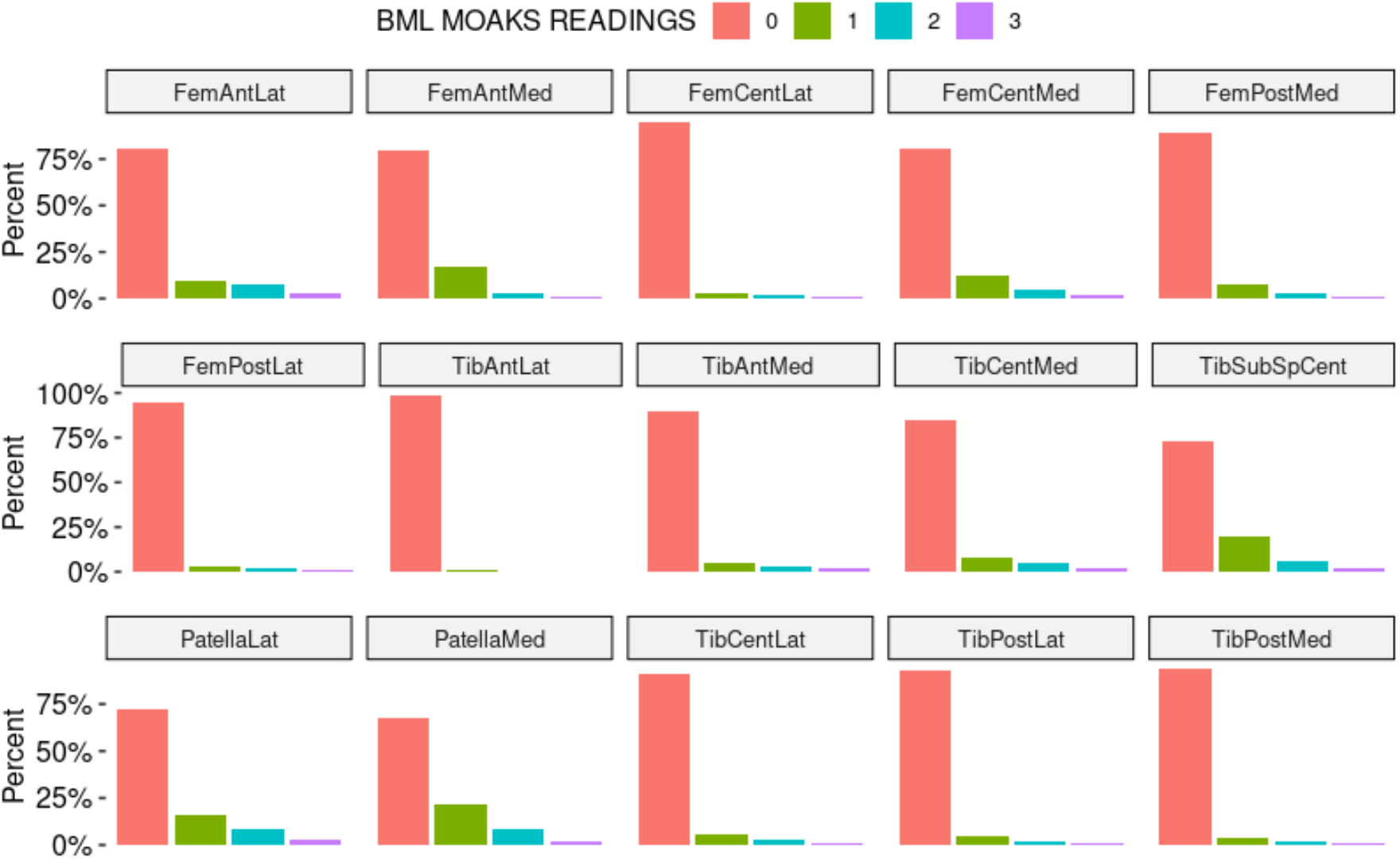
Bar plots of MOAKS scores for each of 15 sub-regions for all data

**Supplementary Figure 2.**
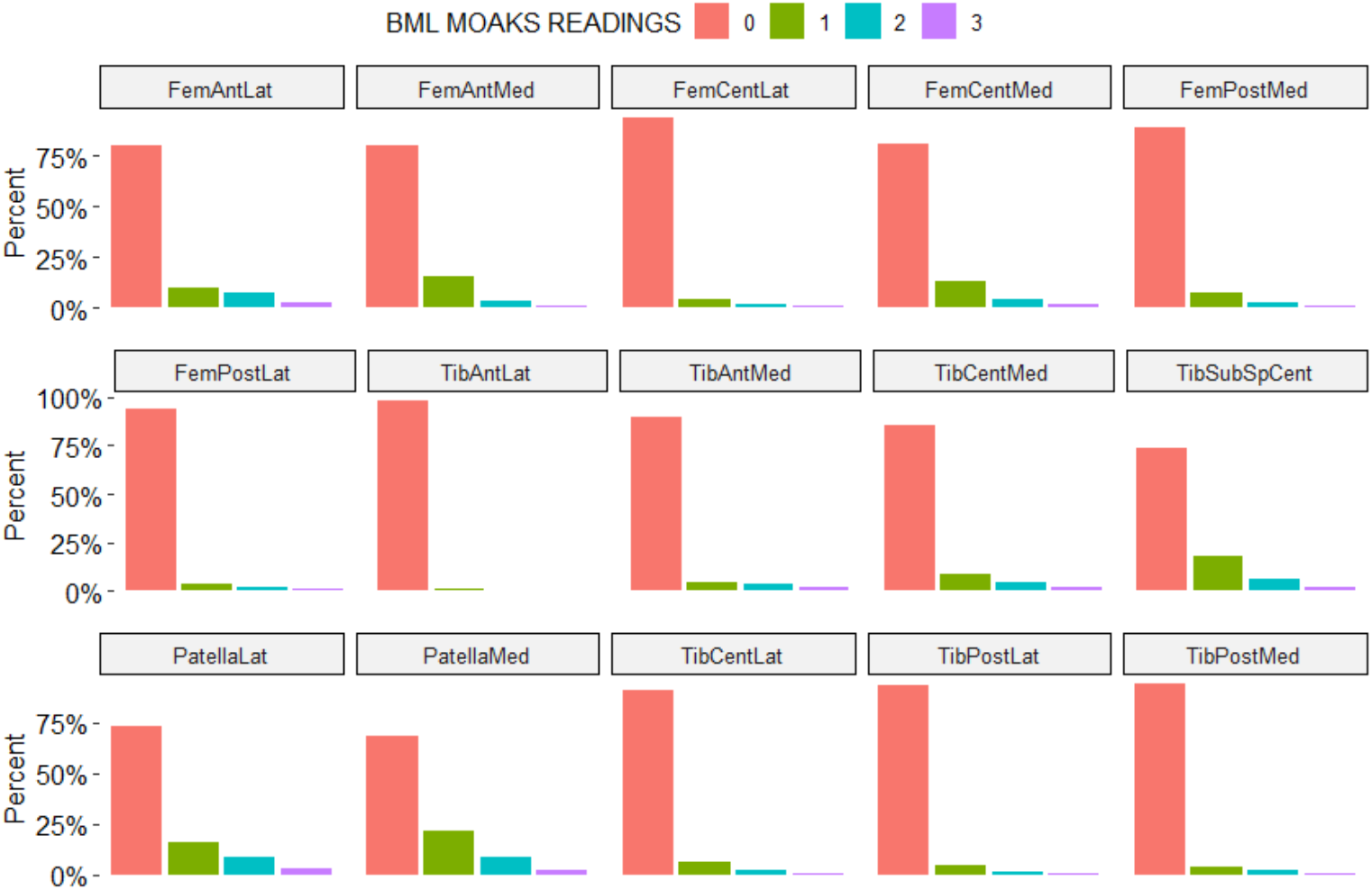
Bar plots of MOAKS scores for each of 15 sub-regions for training data

**Supplementary Figure 3.**
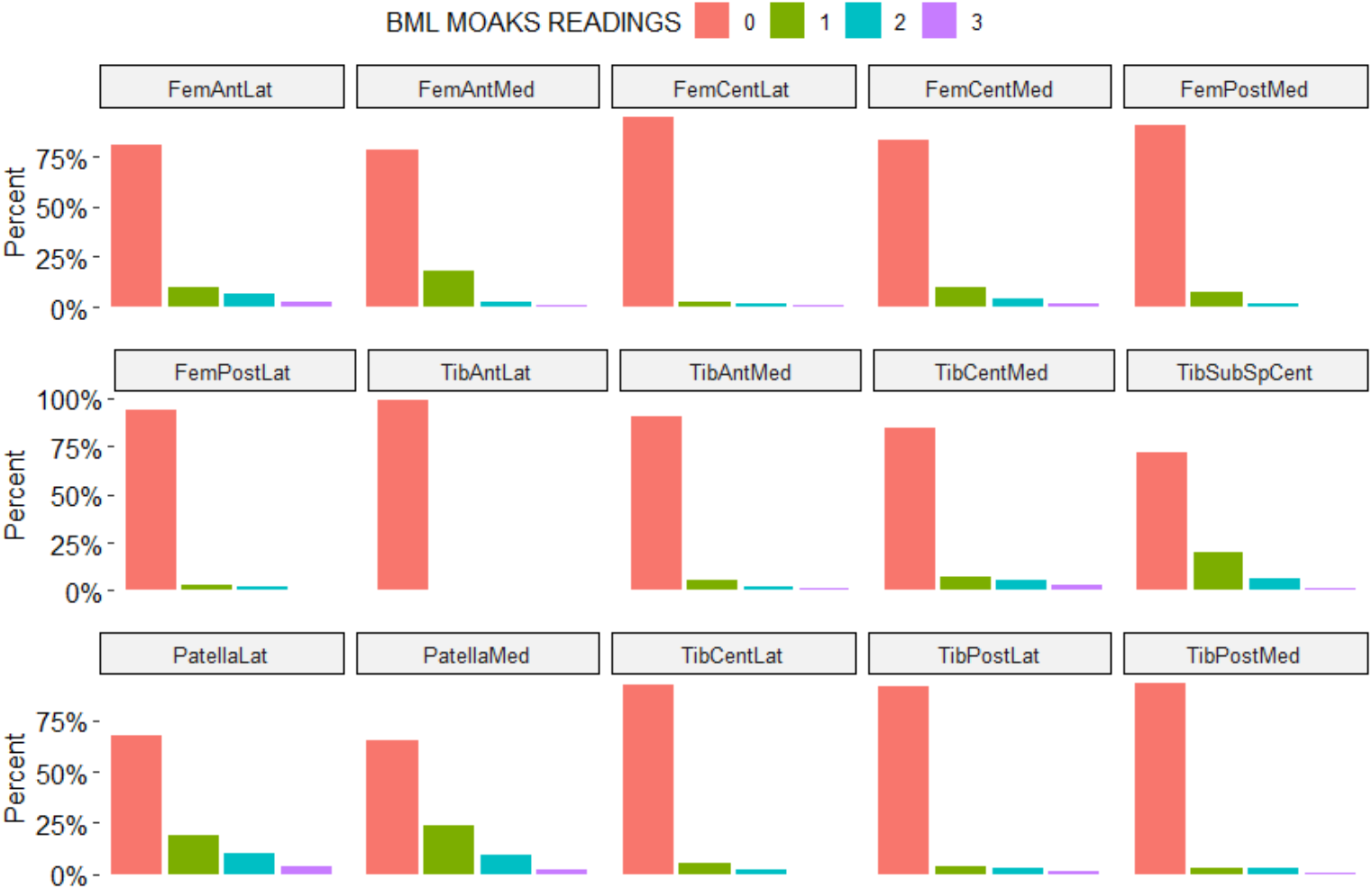
Bar plots of MOAKS scores for each of 15 sub-regions for validation data

**Supplementary Figure 4.**
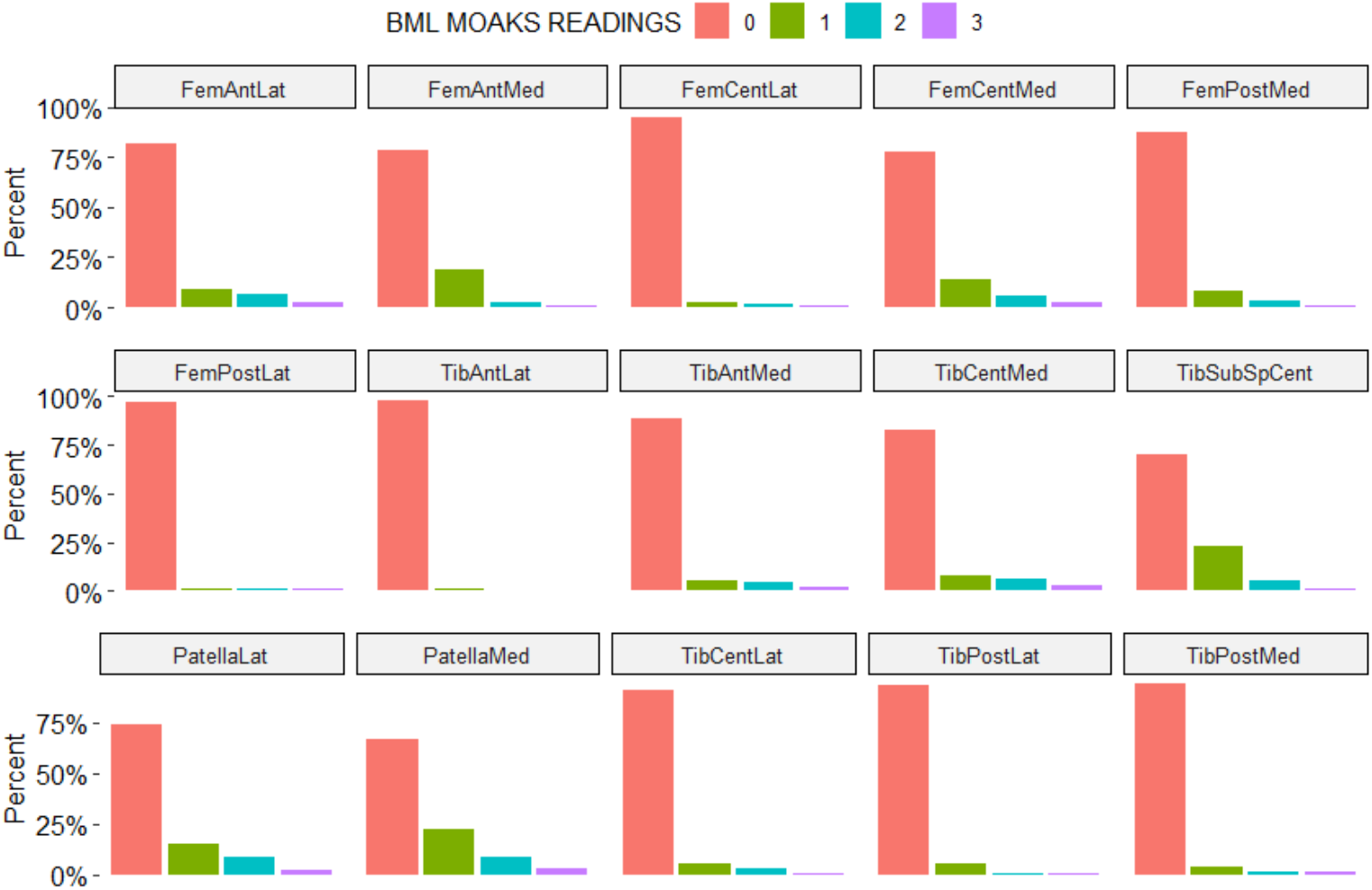
Bar plots of MOAKS scores for each of 15 sub-regions for testing data

